# Systematic review of reviews of symptoms and signs of COVID-19 in children and adolescents

**DOI:** 10.1101/2020.10.16.20213298

**Authors:** Russell Viner, Joseph Ward, Lee Hudson, Melissa Ashe, Sanjay Patel, Dougal Hargreaves, Elizabeth Whittaker

**Author notes:** Correspondence: Prof. Russell Viner, UCL Great Ormond St. Institute of Child Health, 30 Guilford St. London WC1N 1EH.

## Abstract

**Objective:** To undertake a systematic review of reviews of the prevalence of symptoms and signs of COVID-19 in those aged under 20 years?

**Design:** Narrative systematic review of reviews. PubMed, medRxiv, Europe PMC and COVID-19 Living Evidence Database were searched on 9 October 2020.

**Setting:** All settings, including hospitalised and community settings.

**Patients:** CYP under age 20 years with laboratory-proven COVID-19.

**Study review, data extraction and quality:** Potentially eligible articles were reviewed on title and abstract by one reviewer. Quality was assessed using the modified AMSTARS criteria and data were extracted from included studies by two reviewers.

**Main outcome measures:** Prevalence of symptoms and signs of COVID-19

**Results:** 1325 studies were identified and 18 reviews were included. Eight were high quality, 7 medium and 3 low quality. All reviews were dominated by studies of hospitalised children. The proportion who were asymptomatic ranged from 14.6 to 42%. Fever and cough were the commonest symptoms; proportions with fever ranged from 46 to 64.2% and with cough from 32 to 55.9%. All other symptoms or signs including rhinorrhoea, sore throat, headache, fatigue/myalgia and gastrointestinal symptoms including diarrhoea and vomiting are infrequent, occurring in less than 10-20%.

**Conclusions:** Fever and cough are the most common symptoms in CYP with COVID-19, with other symptoms infrequent. Further research on symptoms in community samples are needed to inform pragmatic identification and testing programmes for CYP.

## Background

Most countries rely upon symptom based testing systems for SARS-CoV-2, the virus that causes COVID-19. This is based upon the assumption that testing symptomatic individuals and tracing, testing and isolating their positive contacts, is the most efficient way of using limited testing resources. In the UK the criteria for testing individuals for SARS-CoV-2 are the presence of fever, a new persistent cough and loss or change in smell (https://www.nhs.uk/conditions/coronavirus-covid-19/symptoms/coronavirus-in-children/) with similar symptom-based case definitions operating in other countries.

Such case definitions imply that individuals with fever and/or cough have COVID-19 until proven otherwise, requiring those who are symptomatic to quarantine until a negative SARS-CoV-2 test is obtained. Despite evidence that individual symptoms and signs have relatively poor diagnostic properties for COVID-19 in adults, other causes of fever and cough are uncommon in healthy adults.^1^ However children and young people (CYP), particularly young children, may have 8-10 upper respiratory infections (URI) per year, ^2 3^ particularly over winter. Thus fever and URI symptoms are likely to be less reliable indicators of SARS-COV-2 positivity in children, even when virus prevalence is high.

The resumption of schooling in the UK in September 2020 led to ten-fold increase in numbers of 0-18 year olds reporting potential COVID-19 symptoms in England.^4^ This almost certainly represented large numbers of children attending school with mild fevers and URI symptoms, caused by a variety of common circulating winter viruses Yet these URI symptoms led to many CYP being asked to quarantine or be tested for SARS-CoV-2 before returning to school.^5^ Moreover there has been some uncertainty about which symptoms indicate likely COVID-19 in children and young people (CYP), as some recent studies have suggested that other symptoms such as gastrointestinal symptoms^6^ or fatigue^6 7^ may be common in CYP with COVID-19.

Policy on case definitions and testing policy for COVID-19 in CYP requires evidence on those symptoms which are most commonly associated with test positivity, but also those that are not. In view of a rapidly growing literature in this area including a number of systematic reviews, we undertook a systematic review of reviews (or umbrella review) of symptoms or signs of proven COVID in CYP aged under 20 years.

## Methods

We undertook a rapid systematic review of systematic reviews, reporting Methods and Findings using the PRISMA checklist.^8^

### Review question

Our review question was: What is the prevalence of symptoms and signs of COVID-19 in those aged under 20 years?

### Search strategy

We searched four databases (PubMed; medRxiv; COVID-19 Living Evidence database; European PMC) on 9 October 2020. Search terms in PubMed were (“COVID-19”[Text Word] OR “2019-nCoV”[Text Word] OR “SARS-CoV-2”[Text Word]) AND (“child*”[All Fields] OR “infant*”[All Fields]) AND (“symptom”[All Fields] OR “clinical presentation”[All Fields] OR “Signs”[All Fields]), applying the filters for Meta-Analysis or Systematic review. Search terms in the medRxiv preprint server were: “COVID-19 AND child AND systematic review AND (symptom OR sign OR clinical)” – note that these searches were undertaken separately due to the inability to perform Boolean searches in this database. The COVID-19 Living Evidence database (https://zika.ispm.unibe.ch/assets/data/pub/search_beta/) and European PMC (https://europepmc.org) were searched subsequently using similar terms. We did not limit studies by date or language. Identified relevant reviews were hand-searched for additional likely studies. Studies were also identified through the professional networks of the authors.

### Eligibility

We included systematic reviews or meta-analyses of clinical presentations (symptoms or signs) of laboratory-proven COVID-19 disease in CYP. We only included reviews which:

i. Systematically searched and reviewed the literature using prespecified protocols
ii. examined CYP from 0 – 19 years. Studies with a wider age range which provided data on CYP separately were eligible
iii. included only studies with laboratory-proven SARS-CoV-2 cases
iv. assessed and reported the frequency or prevalence of symptoms or signs of COVID-19

We excluded studies of investigation results, cross-age studies that did not separate data on CYP, studies of conditions linked with COVID-19 e.g. (Paediatric Inflammatory Multisystem Syndrome (PIMS), and preprints that had been deposited for >5 months without update or publication.

### Study selection

The search and inclusion flowchart is shown in Figure 1. Titles and abstracts were reviewed and potentially eligible articles identified by one reviewer (RV). Full text review and agreement on final inclusion was undertaken by two reviewers (RV; JW). The abstracts of 1325 studies were reviewed and 21 potentially eligible articles were identified for full text review, with 1 additional study identified through handsearching of citation lists and 3 studies through professional networks. After further review, 18 reviews are included in this review.

**Figure 1.**
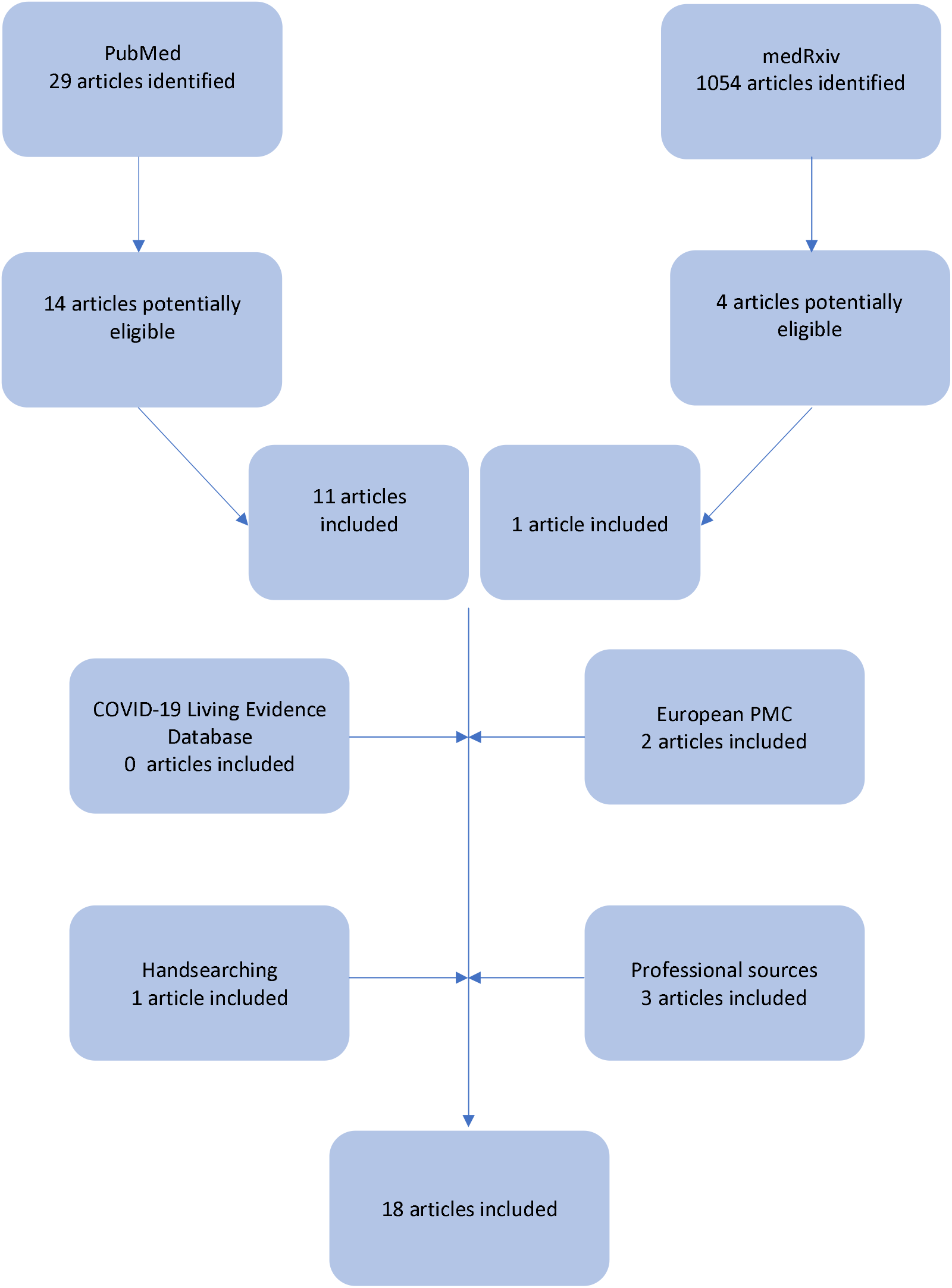
PRISMA flow chart

### Data extraction

Descriptive findings were extracted to a spreadsheet and checked for accuracy by two reviewers (JW, RV).

### Quality evaluation

Quality including risk of bias was assessed using an adapted version of Assessing the Methodological Quality of Systematic Reviews (AMSTARS).^9 10^ We characterised reviews as high, medium or low quality. High-quality reviews were required to have provided a priori designs; searched at least two bibliographic databases; searched for reports regardless of publication type; listed and described included studies; used at least two people for data extraction; documented the size and quality of included studies and used this to inform their syntheses; synthesised study findings narratively or statistically; assessed the likelihood of publication bias; and included a conflict of interest statement. Those unable to meet 6 of these 8 requirements were characterised as low quality, with those meeting 6 or 7 criteria characterised as medium quality. Note we made no attempt to assess the quality of studies included in each review. Quality assessments for each review are shown in Appendix Table 1.

**Table 1.** Characteristics of included studies

| Study | Source | Type | Context | Search end | Age | Case definition | Included studies | Total children | Countries of studies | Quality summary |
| --- | --- | --- | --- | --- | --- | --- | --- | --- | --- | --- |
| Mantovani et al.<sup>11</sup> | Professional | SR/MA | All | 11 April | NS | NS | 19 | 2855 | China 17, USA, Spain | High |
| De Sousa et al.<sup>12</sup> | PubMed | SR | All | 6 April | <18y | RT-PCR+ | 38 | 1124 | China 33, Italy, Iran, Singapore, South Korea, Vietnam | Low |
| Yasuhara et al.<sup>13</sup> | PubMed | SR | All | 20 June | <18y | RT-PCR+ | 46 | 114 | China 20, USA 13, 2 Italy, 2 Iran, Korea, Vietnam, Malaysia, Spain, Lebanon, Belgium, UK, Turkey, France | Low |
| Hoang et al.<sup>14</sup> | PubMed | SR | All | 14 May | ≤21y | RT-PCR+ | 131 | 7780 | 26 countries; China made up 64% of studies | High |
| Cui et al.<sup>15</sup> | PubMed | SR/MA | All | 30 April | <18y | RT-PCR+ | 48 | 5829 | China 43, Singapore, Korea, Spain, USA, Iran | High |
| Raba et al.<sup>16</sup> | Professional | SR | All; under 1 year | 7 April | <1y | RT-PCR+ | 18 | 160 | China 17, Vietnam | Medium |
| Zhang et al.<sup>17</sup> | PubMed | SR/MA | All | 4 May | 0-18y | 'laboratory confirmed COVID-19' | 46 | 551 | China 35, Iran 2, Italy 3, South Korea, Malaysia, Singapore, Spain, USA, Vietnam | High |
| Ma et al.<sup>18</sup> | PubMed | SR/MA | Hospitalised | 21 April | NS | RT-PCR+ | 15 | 486 | China | Medium |
| Ding et al.<sup>19</sup> | Professional | SR/MA | All | 1 April | 0-17y | RT-PCR+ | 14 | 371 | China | High |
| Christophers et al.<sup>20</sup> | PubMed | SR | All | 15 May | 1 mnth-18y | RT-PCR+ | 21 | 123 | China 16, USA, Italy 2, Iran, Malaysia, | Medium |
| Liguoro et al.<sup>21</sup> | PubMed | SR | All | 1 May | 0-18y | 'confirmed cases' | 62 | 7480 | China 49, South Korea 3, Italy 5, Vietnam, Iran, USA, Spain, Malaysia | Medium |
| Assaker et al.<sup>22</sup> | PubMed | SR/MA | All | 3 May | NS | RT-PCR+ | 28 | 1614 | China 24, Malaysia, Spain, Italy, USA | Medium |
| Chang et al.<sup>23</sup> | Handsearch | SR/MA | All | 15 March | <18y | RT-PCR+ | 9* | 93 | China | Medium |
| Wang et al.<sup>24</sup> | medRxiv | SR/MA | All | 31 | <18y | WHO | 49<sup>+</sup> | 1667 | 45 China 1 Singapore, | High |
|  |  |  |  | March |  | definition |  |  | 1 Korea, 1 Vietnam, 1 Iran |  |
| Jahangir et al.<sup>25</sup> | PubMed | SR | All | 9 April | 0-19y | NS | 27 | NS | China, South Korea, Spain, USA: numbers not stated | Low |
| Pei et al.<sup>26</sup> | PubMed | SR | All | 3 March | 0-18y | RT-PCR+ | 5 | 70 | China | Medium |
| Kharoud et al.<sup>27</sup> | Europe PMC | SR/MA | All | 15 July | 0-18y | 'laboratory confirmed COVID-19' | 37 | 668 | China 33, Italy 2, USA, Spain | High |
| Wang, Cui et al.<sup>28</sup> | Europe PMC | SR/MA | Gastrointestinal symptoms only | 10 August | NS | NS | 38 | 3028 | China 21, other countries (not listed) 17 | High |

Notes:
SR: systematic review
MA: meta-analysis
Contact: All = all contexts;
NS: not stated
Search end: all dates are in 2020.
Countries: where countries have no number next to them, each country had 1 included study
\*In Chang et al., 2 studies examined neonates only which were not included in the meta-analyses
+In Wang et al. only studies with at least 9 cases were included in the meta-analyse

### Data synthesis and summary measures

We confined our analysis to descriptive measures of the prevalence of each symptom or sign in each included review. We made no attempt to quantitatively summarise findings across reviews.

## Results

We included 18 reviews in our analysis.^11-28^ Characteristics of the included reviews are shown in Table 1; all are freely available online (see Appendix Table 2). All reviews were in English. The great majority of studies included in all reviews were from China, with studies also from Italy, Spain, South Korea, Malaysia, Singapore, Vietnam, Iran and the USA. Eight were high quality,^11 14 15 17 19 24 27 28^ 7 were medium, ^16 18 20-23 26^ and 3 were low quality.^12 13 25^ The very great majority of studies within the reviews were of hospitalised CYP. No reviews specifically focused on symptoms or signs amongst community samples of infected children, and none compared symptoms in hospitalised compared with non-hospitalised children. One review included only studies of infants^16^, one included a comparison of symptoms in children with those in adults from China,^26^ and one examined only gastrointestinal symptoms.^28^ One study provided descriptive data from included studies only and did not report pooled estimates of symptom prevalence.^25^

Figure 2 shows the proportions of CYP in each review with reported symptoms or signs of COVID-19 data shown by study in Appendix Table 3). The proportion who were asymptomatic ranged from 14.6 to 42%. Fever and cough were the commonest symptoms; proportions with fever ranged from 46 to 64.2% and with cough from 32 to 55.9%. All other symptoms or signs were present at less than 10-20%. Vomiting, diarrhoea and abdominal pain were reported separately in the majority of studies, however some reported gastrointestinal symptoms together. These estimates ranged from 7.4%^19^ to 17.7%.^28^ All studies which reported fatigue and myalgia did so as ‘fatigue or myalgia’, with the exception of Assaker et al.^22^ who reported myalgia in 14% and fatigue in 8%.

**Figure 2.**
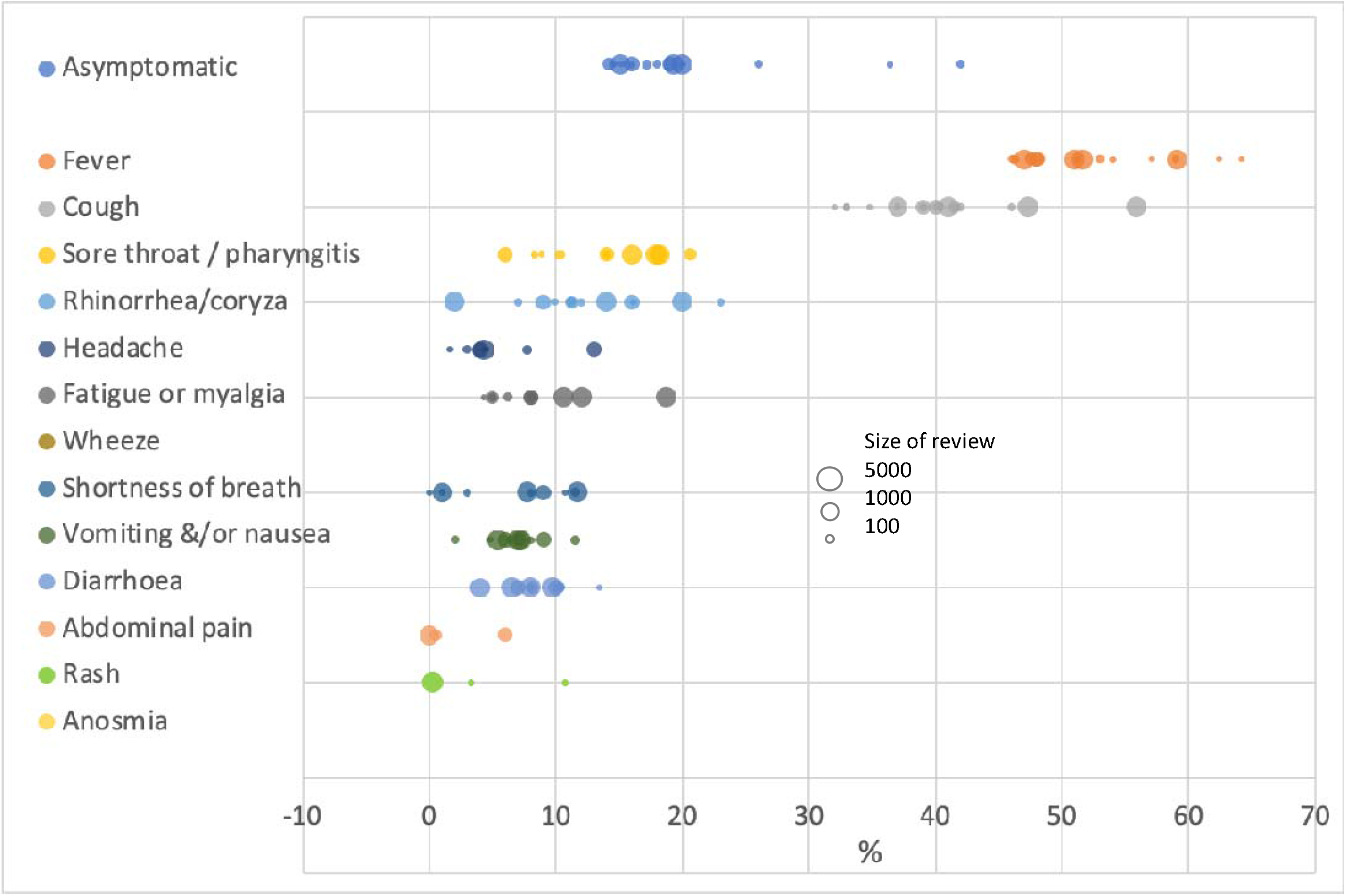
Proportions with symptoms and signs across reviews

Only two studies reported on combinations of symptoms or signs. Wang et al.^24^ in a high quality review reported that 30% had both fever and cough, whilst a medium quality review by Christophers et al.^20^ reported that 58.4% had only a single symptom; in 23.2% this was fever, 5.6% cough and in 1.6% diarrhoea.

Few studies addressed differences in symptom profiles by age amongst CYP. A large high quality review by Cui et al.^15^ reported that proportions with fever and cough were similar in infants to those in older children, conclusions supported by a small low quality review by Yasuhara et al.^13^

In contrast, a small medium quality review, Christophers et al.^20^ reported that fever appeared more common in older children (65%) than in infants (48%). Christophers et al.^20^ also reported that vomiting and diarrhoea occurred largely in those under 9 years, whereas Yasuhara et al.^13^ reported gastrointestinal symptoms to be more common in adolescents, although numbers were small and symptoms uncommon in all ages.

Only one review compared CYP with adults. Pei et al.^26^ undertook a medium quality review comparing symptoms in 170 children and 275 adults, restricting the analysis to studies from China. They reported that that CYP were more likely to be asymptomatic than adults (20% compared with 5.5%).

## Discussion

We identified 18 systematic reviews and meta-analyses that were informative about the prevalence of symptoms and signs of COVID-19 in CYP. Findings show clearly that fever and cough are the most common symptoms occurring in around 40-60% of infected CYP, with all other symptoms occurring at much lower prevalences, under 10-20%. Common URI symptoms such as rhinorrhoea or sore throat are uncommon in COVID-19, as are somatic symptoms such as headache and fatigue/myalgia and gastrointestinal symptoms including diarrhoea and vomiting. We found no data on the frequency of loss or change in smell in these reviews.

Few data were available on the frequency of combinations of symptoms although one study reported that the combination of fever and cough occurred in around one-third of CYP, less common that fever or cough alone. The common symptoms of fever and cough appeared equally prevalent across younger and older children. There were insufficient data to conclude that there were age-differences in the frequency of other symptoms between younger and older children.

Findings from this study are consistent with recently published large UK^29^ and pan-European^30^ studies of children hospitalised with COVID-19, both of which reported fever occurring in approximately 65%. The UK study of over 650 CYP hospitalised with COVID-19 found similarly low proportions with symptoms other than fever and cough.

The main limitation of this review is that the great majority of studies in the reviews we examined were of hospitalised children. Whilst early in the pandemic all positive children were hospitalised in some countries regardless of severity, the inclusion of largely hospitalised children is likely to strongly bias these findings towards those with clinical disease and more severe disease. This is likely to inflate the prevalence of symptoms and decrease estimates of the proportion who are asymptomatic. Other studies have suggested that only around 20%^31^ -50%^6^ of population samples of CYP have symptoms. Findings from the small number of available community samples suggest that lower proportions present with fever (30%^6 32^ -40%^33^) and cough,(10%^6^ -35%)^32 33^ consistent with a higher proportion who are asymptomatic.

Our findings are subject to a number of other limitations. Some studies were included in multiple reviews in our study. For this reason we did not undertake a meta-summary of findings across reviews. However even in the narrative review undertaken here duplication of studies may still inflate the importance of certain studies compared with others. There was considerable heterogeneity across the included reviews and the risk of publication bias varied by symptom in some reviews. The majority of reviews included data from the early months of the pandemic, and only 3 reviews included studies published after June 2020.^13 27 28^ This leads to the domination of findings by studies from China and by case series of hospitalised children with more severe COVID-19, as these studies were more quickly set up early in the pandemic. One review was excluded as it was judged not to be a systematic review.^34^ A review of COVID-19 in CYP with developmental disabilities was excluded as it contained no useable data.^35^

## Conclusions

Available data suggests that fever and cough are the predominant symptoms of COVID-19 in CYP, supporting their use in case definitions of potential COVID-19 in this age-group. Other symptoms are uncommon and their inclusion in case definitions and testing systems for COVID-19 would lower specificity markedly. Rhinorrhoea and sore throat are infrequent in COVID-19 in CYP, which, given the frequency of URI and associated symptoms in young children, suggests that their presence is much more likely to indicate infection with viruses other than SARS-CoV-2. Further data on symptoms in community samples including in schools are urgently needed to inform pragmatic identification and testing programmes for CYP and reduce misclassification of CYP as potential COVID-19 cases requiring isolation of peers and families.

## What is already known on this topic

- Symptom-based case definitions for likely COVID-19 may misclassify children as young children may have 8-10 upper respiratory infections per year.
- Some recent studies have suggested that gastrointestinal symptoms or fatigue maybe more prevalent amongst children than cough and fever
- Policy on case definitions and testing policy for COVID-19 in CYP requires evidence on those symptoms which are most commonly associated with test positivity, but also those that are not. In

## What this study adds

- Fever and cough are the most common symptoms of COVID-19 in children and young people, by some margin.
- Other symptoms, including rhinorrhoea, sore throat and diarrhoea and vomiting, are infrequent. Their inclusion in symptom profiles for COVID-19 is likely to markedly reduce specificity

## Supporting information

PRISMA checklist

## Data Availability

All included articles in this systematic review are available open-access on-line.

## Author contribution

RV, EW, SP and MW conceptualised the paper. RV undertook the searching of databases and identification of potentially eligible articles. JW and RV agreed the inclusion of the final articles, assessed quality and extracted data. All authors contributed to editing of the final paper.

