## Appendix for "Systematic review of reviews of symptoms and signs of COVID-19 in children and adolescents"

### Appendix Table1. Quality summary of included studies

| STUDY | **provides a priori design including RQ and inclusion criteria** | **duplicate searching and data extraction** | **comprehensive literature review (≥2 databases)** | **listed and described included studies** | **Quality of included studies assessed** | **synthesized narratively or statistically** | **assessed the likelihood of publication bias** | **mention conflict of interest** | **Summary** |
| --- | --- | --- | --- | --- | --- | --- | --- | --- | --- |
| Mantovani | Y | Y | Y | Y | Y | Y | Y | Y | High |
| De Souza | Y | Y | N | Y | N | Y | N | Y | Low |
| Yasuhara | Y | Y | Y | Y | N | Y | N | Y | Low |
| Hoang | Y | Y | Y | Y | Y | Y | Y | Y | High |
| Cui | Y | Y | Y | Y | Y | Y | Y | Y | High |
| Raba | Y | Y | Y | Y | Y | Y | N | Y | Medium |
| Zhang | Y | Y | Y | Y | Y | Y | Y | Y | High |
| Ma | Y | Y | Y | Y | Y | Y | N | Y | Medium |
| Ding | Y | Y | Y | Y | Y | Y | Y | Y | High |
| Christophers | Y | Y | Y | Y | Y | Y | N | Y | Medium |
| Liguoro | Y | Y | Y | Y | Y | Y | N | Y | Medium |
| Assaker | Y | Y | Y | N | Y | Y | Y | Y | Medium |
| Chang | Y | Y | Y | N | Y | Y | Y | Y | Medium |
| Wang | Y | Y | Y | Y | Y | Y | Y | Y | High |
| Jahangir | N | Y | N | Y | N | Y | N | Y | Low |
| Pei | Y | Y | Y | Y | Y | Y | Y | N | Medium |
| Kharoud | Y | Y | Y | Y | Y | Y | Y | Y | High |
| Wang Cui | Y | Y | Y | Y | Y | Y | Y | Y | High |

### Appendix Table 2. Web references for included studies

1. Mantovani <https://www.nature.com/articles/s41390-020-1015-2>
2. De Sousa <https://onlinelibrary.wiley.com/doi/full/10.1002/ppul.24885>
3. Yasuhara <https://onlinelibrary.wiley.com/doi/epdf/10.1002/ppul.24991>
4. Hoang <https://www.ncbi.nlm.nih.gov/pmc/articles/PMC7318942/>
5. Cui <https://www.ncbi.nlm.nih.gov/pmc/articles/PMC7436402/pdf/JMV-9999-na.pdf>
6. Raba <https://onlinelibrary.wiley.com/doi/10.1111/apa.15422>
7. Zhang <https://www.ncbi.nlm.nih.gov/pmc/articles/PMC7300763/>
8. Ma <https://www.ncbi.nlm.nih.gov/pmc/articles/PMC7323441/pdf/JMV-9999-na.pdf>
9. Ding <https://www.ncbi.nlm.nih.gov/pmc/articles/PMC7350605/pdf/fped-08-00431.pdf>
10. Christophers <https://www.nature.com/articles/s41390-020-01161-3>
11. Liguoro <https://www.ncbi.nlm.nih.gov/pmc/articles/PMC7234446/>
12. Assaker <https://www.ncbi.nlm.nih.gov/pmc/articles/PMC7261471/>
13. Chang <https://www.ncbi.nlm.nih.gov/pmc/articles/PMC7161491/>
14. Wang <http://atm.amegroups.com/article/view/42970/html>
15. Jahangir <https://www.hkmj.org/earlyrelease/hkmj208646.htm>
16. Pei	<https://www.ncbi.nlm.nih.gov/pmc/articles/PMC7357530/>
17. Kharoud	<https://www.medrxiv.org/content/10.1101/2020.09.23.20200410v1>
18. Wang Cui <https://www.researchsquare.com/article/rs-34733/v2>

### Appendix Table 3. Proportions with symptoms and signs by review

|  | Mantovani | De Sousa | Yasuhara | Hoang | Cui | Raba | Zhang | Ma | Ding | Christophers | Liguoro | Assaker | Chang | Wang | Pei | Kharoud | Wang Cui |
| --- | --- | --- | --- | --- | --- | --- | --- | --- | --- | --- | --- | --- | --- | --- | --- | --- | --- |
| Asymptomatic |  | 14.2 | 15.2 | 19.3 | 20 | 16 | 18 | 42 | 36.4 | 14.6 | 15.1 | 16 | 26 | 19 | 20.0 | 17.2 |  |
| Fever | 47 | 47.5 | 64.2 | 59.1 | 51 | 54 | 53 | 46 | 51.2 | 62.4 | 51.6 | 48 | 59 | 48 | 57.1 | 46.2 |  |
| Cough | 37 | 41.5 | 34.8 | 55.9 | 41 | 33 | 39 | 42 | 37 | 32 | 47.3 | 40 | 46 | 39 | 32.9 | 40.2 |  |
| Sore throat / pharyngitis |  | 20.6 | 8.9 | 18.2 | 16 |  | 14 |  | 8.3 |  | 17.9 | 14 |  | 6 |  | 10.3 |  |
| Rhinorrhea/coryza | 2 | 11.2 | 16.1 | 20 | 14 | 23 | 7 | 12 | 9.9 |  |  | 16 |  | 9 |  | 11.2 |  |
| Headache |  |  | 4.5 | 4.3 |  |  | 3 |  |  | 1.6 |  | 13 |  | 4 | 4.3 | 7.7 |  |
| Fatigue or myalgia |  | 5 |  | 18.7 | 12 |  | 5 | 8 |  |  | 10.6 | 8 |  | 8 | 4.3 | 6.23 |  |
| Wheeze |  |  |  |  |  |  |  |  |  |  |  |  |  |  |  |  |  |
| Shortness of breath | 1 |  | 10.7 | 11.7 |  | 3 | 8 |  | 1 |  | 7.7 |  |  | 9 | 0 | 11.5 |  |
| Vomiting &/or nausea |  | 7.1 | 6.3 | 5.4 | 7 |  | 2 | 8 |  | 4.8 | 7.2 | 9 |  | 6 |  | 11.5 |  |
| Diarrhoea | 4 | 8.1 | 13.4 | 6.5 | 8 |  | 8 | 10 |  | 10.4 | 9.7 | 10 |  | 7 |  | 10.3 |  |
| Abdominal pain | 0 | 0.5 |  |  |  |  |  |  |  |  |  | 6 |  |  |  |  |  |
| Any gastrointestinal symptom (excluding above) |  |  |  |  |  | 16 |  |  | 7.4 |  |  |  | 12 |  | 7.1 |  | 17.7 |
| Rash |  |  | 10.7 | 0.25 |  |  |  |  |  | 3.3 |  |  |  |  |  |  |  |
